# Temporal trends and practice variation of paediatric diagnostic tests in primary care

**DOI:** 10.1101/2024.05.20.24307611

**Authors:** Elizabeth T Thomas, Diana R Withrow, Peter J Gill, Rafael Perera, Carl Heneghan

## Abstract

**Objective:** The primary objective was to investigate temporal trends and between-practice variability of paediatric test use in primary care.

**Methods and analysis:** This was a descriptive study of population-based data from primary care consultation records from January 1, 2007, to December 31, 2019. Children aged 0 to 15 who were registered to one of the 1,464 practices and had a diagnostic test code in their clinical record were included. The primary outcome measures were: 1) temporal changes in test rates measured by the average annual percent change (AAPC), stratified by test type, gender, age group, and deprivation level and 2) practice variability in test use, measured by the coefficient of variation (CoV).

**Results:** 14,299,598 diagnostic tests were requested over 27.8 million child-years of observation for 2,542,101 children. Overall test use increased by 3.6%/year (95% CI 3.4 to 3.8%) from 399/1,000-child-years to 608/1,000 child-years, driven by increases in blood tests (8.0%/year, 95% CI 7.7 to 8.4), females aged 11-15 (4.0%/year, 95% CI 3.7 to 4.3), and the most socioeconomically deprived group (4.4%/year, 95% CI 4.1 to 4.8). Tests subject to the greatest temporal increases were fecal calprotectin, fractional exhaled nitric oxide (FeNO), and vitamin D. Tests classified as high use and high practice variability were iron studies, vitamin D, vitamin B12, folate, and coeliac testing.

**Conclusions:** In this first nationwide study of paediatric test use in primary care, we observed significant temporal increases and practice variability in testing. This reflects inconsistency in practice and diagnosis rates, and a scarcity of evidence-based guidance. Increased test use generates more clinical activity with significant resource implications, but conversely may improve clinical outcomes. Future research should evaluate whether increased test use and variability is warranted by exploring test indications and test results, and directly examine how increased test use impacts on quality of care.

**Key Messages:** *What is already known on this topic:* Previous research has shown that test use in adults within UK primary care sharply increased since 2000 and that there is a high degree of practice variation in test use. To date, no population-based studies have analysed paediatric test use in this setting.

*What this study adds:* In England between 2007 and 2019, diagnostic test use increased by 4% per year, from 399 tests/1,000 child-years to 608 tests/1000-child years. Test increases were driven blood tests, especially in females aged 11-15 years of age, and children in the most deprived socioeconomic group. Specific tests that increased by the greatest margin include faecal calprotectin, fractional exhaled nitric oxide (FeNO), and vitamin D testing. Tests subject to the greatest practice variation by 2019 were FeNO, hearing tests, and vitamin D levels.

*How this study might affect research, practice or policy:* Variability in test use highlights a lack of standardised guidance and evidence in pediatric diagnostics, which has significant implications for downstream diagnostic activity, treatment, referrals and healthcare costs.

## Introduction

It has been reported that 70% of clinical decisions involve the use of diagnostic tests.[1] Paediatric test use has not previously been characterised in large-scale population studies, especially in primary care, where most paediatric health contacts occur. Compared with adults, children more commonly present with undifferentiated symptoms, and non-specific complaints such as abdominal pain, headaches, and fatigue often have no identifiable underlying cause.[2] While tests are one of the many diagnostic strategies available to clinicians, they must balance the risks of over-investigation and unnecessary referrals with missing or delaying a diagnosis. It is difficult to achieve the right balance of care, and the threshold to test, treat or refer varies among clinicians [3], which can lead to substantial variation in the health care delivered to children.[4, 5]

Measuring variation in testing can identify tests that are potentially overused or underused, with both over and under use having potentially harmful consequences.[6] For the patient, overuse can result in testing cascades, which cause harm through unnecessary treatment, physical, and emotional trauma. [7] For the health care system, overuse generates additional workload for clinicians and can lead to unnecessary referrals, healthcare contacts, and health spending in an already overburdened system.[8, 9] Underuse of tests can result in missed or delayed diagnosis and treatment with potentially serious physical, emotional, and financial consequences for patients, families, and clinicians.

Studies analysing test use in adults have reported that test use in primary care increased by 8.5% annually[10, 11]. We previously published a study that described temporal trends in paediatric blood tests in Oxfordshire from 2005 to 2019 and found that test use increased in outpatient (specialty and general practice) settings compared with inpatient services where test rates remained stable. [12] The overall aim of this study was therefore to quantify and analyse temporal change and variation in paediatric diagnostic tests in primary care across England. The specific objectives were to 1) quantify how paediatric diagnostic test use has changed over time and varied by primary care practice and 2) determine how demographic and socioeconomic factors impact test use.

## Methods

### Study design and sample

This was a retrospective population-based observational study using routinely collected data from the electronic health records of children aged 0 – 15 years presenting to primary care practices in the UK (99% from England, 1% from Northern Ireland) from 1 January 2007 to 31 December 2019.[13] The UK National Health Service is a publicly funded healthcare system, where primary care practitioners are gatekeepers to specialist paediatric care and carry out most health care consultations for children.[14] Person-years were estimated as the time from birth or registration date, until 16 years of age, death, end of the study period, or transfer out of the practice.

### Data Source

The Clinical Practice Research Datalink (CPRD) Aurum contains routinely collected data from primary care practices that use EMIS Web® electronic patient record system software.[15] The data encompass 19.9% of the UK population and 16.6% of UK primary care practices.[13] To gather information on socioeconomic status, we obtained linked data for practice-level index of multiple deprivation (IMD), which is a composite measure derived from indicators for the following domains related to deprivation: income, employment, education and skills, health, housing, crime, access to services, and living environment.[16] CPRD data is quality-assured, has been shown to be representative of the national UK population due to its breadth and coverage, and has been extensively validated for use in observational research.[17]

### Included tests

For overall metrics of paediatric test use, we included all diagnostic tests, including blood tests, imaging, physiological tests, and invasive procedures such as colonoscopy. Physical examination findings, anthropometric measurements, and vital signs were excluded. When analysing trends and variation in specific tests, we used a subset of tests restricted to (1) the 25 most frequently requested tests during the study period (2) tests that are frequently reported by primary care providers as requested for children or perceived to be subject to substantial variation in their use [18], or (3) from other literature that focused on paediatric diagnostic test use in primary care [12, 19, 20]. The resulting 35 included tests comprised approximately 80% of the total tests conducted (see **Appendix Table 1**). Tests were grouped by type: blood tests, imaging, and miscellaneous non-serum laboratory tests.

### Statistical analysis

Crude rates of test use were estimated per 1000 child-years. Age-adjusted annual rates were standardised to the 2019 age distribution.

#### Temporal variation

We used Joinpoint regression to model temporal changes in age-adjusted test rates from 2007 to 2019, which has previously been used in similar studies analysing temporal trends in test use.[10, 12] Points where significant changes in rates occurred (called joinpoints) were identified, and annual percentage changes (APC) between joinpoints were estimated. The joinpoint regression model also provided an estimate of the average annual percentage change (AAPC), a summary measure of the trend from 2007 to 2019, along with the associated p values. Age-adjusted rates, APCs, and AAPCs were stratified by test type, gender, age group, and index of multiple deprivation (IMD) quintile, where 1 represented the least deprived group.

#### Practice variation

Crude rates for practice variation used, as the denominator, child-years contributed by the practice in 2019, where each child contributed the full or partial years they were registered. We estimated an unadjusted coefficient of variation (CoV) by dividing the standard deviation of the unadjusted test rates by the mean. Rates of test use were by practice were adjusted using a generalised linear model with Poisson errors to account for gender (proportion of females), median age of the study population, and deprivation index (IMD decile).[11] Adjusted rates were used to calculate the adjusted CoV.

APCs and AAPCs were modelled in Joinpoint Regression software version 5.0.2.[21] Data cleaning, management and all other analyses were performed using R version 4.3.1.[22]

### Patient and Public Involvement

The patient and public advisory group consisted of three parents who were involved in the planning and design of this study, including consideration of which tests to include for the test-specific analyses. They were also involved in discussions on future directions of this research and dissemination strategies.

## Results

### Characteristics of included participants and tests

There were 14,299,598 tests performed over 27,809,957 child-years of observation from 1 January 2007 to 31 December 2019, among 2,542,101 children of whom 50.4% (1,282,072 of) were females, see **Table 1**. 54.5% of the total tests (7,794,755 of 14,299,598 tests) were performed in females. Blood tests were the most frequently performed type of diagnostic tests (50.1%). Nearly 40% of tests (39.2%) were conducted for children aged 11-15. Overall, the median number of tests per child per year was 2 (Interquartile range [IQR] 1 to 3). Once stratified by age group, the median number of tests per year was 1 for all age groups under 11 and 2 (IQR 1 to 5) for children aged 11-15 years. Patients in more deprived practices were over-represented relative to population deciles.

**Table 1.** Characteristics of included participants and tests.

|  | **Number of tests** | **%** |
| --- | --- | --- |
| **Total** | 14,299,598 | 100.0 |
| **Type of test** |  |  |
| Blood | 7,157,882 | 50.1 |
| Imaging | 885,709 | 6.2 |
| Miscellaneous* | 6,256,007 | 43.7 |
| **Gender** |  |  |
| Male | 6,504,010 | 45.5 |
| Female | 7,794,755 | 54.5 |
| Indeterminate | 833 | 0.0 |
| **Age group** |  |  |
| Under 1 | 1,460,767 | 10.2 |
| 1-5 | 3,186,435 | 22.3 |
| 6-10 | 4,046,845 | 28.3 |
| 11-15 | 5,605,551 | 39.2 |
| **IMD quintile** |  |  |
| 1 (least deprived) | 2,223,937 | 15.6 |
| 2 | 2,133,884 | 14.9 |
| 3 | 2,804,071 | 19.6 |
| 4 | 3,230,863 | 22.6 |
| 5 (most deprived) | 3,906,843 | 27.3 |
|  | **Median tests per person per year** | **IQR** |
| **Overall** | 2 | 1,3 |
| **Gender** |  |  |
| Male | 1 | 1,3 |
| Female | 2 | 1,3 |
| **Age group** |  |  |
| Under 1 | 1 | 1,2 |
| 1-5 | 1 | 1,3 |
| 6-10 | 1 | 1,3 |
| 11-15 | 2 | 1,5 |

Miscellaneous tests include laboratory analysis of non-serum samples (e.g., urine, stool) and physiological measurements.

### Temporal trends in overall test use

The age-adjusted rate of total test use increased from 399 tests per 1,000 child years in 2007 to 608 tests per 1,000 child years in 2019, an average annual percentage increase of 3.6% per year (AAPC 95% CI 3.4 to 3.8%), see **Figure 1a**, **Appendix Table 1**. Test rates initially increased by 5.1% per year (APC 95% CI 4.7 to 5.6%) between 2007 to 2014, then increased by 1.6% per year (APC 95% CI 0.9 to 2.1%) between 2014 and 2019. **Figure 1b** shows temporal changes in test use stratified by test type. The greatest increase was observed for blood tests which increased by 8.0% per year (AAPC 95% 7.7 to 8.4%). Rates of test use by gender and age group are shown in **Figure 1c** and **1d**. Test rates were consistently higher in females compared with males. When stratified by age group, the rates of change were similar for both genders and age groups, except for 11-15-year-olds, where testing increased by 4.0% per year (AAPC 95% CI 3.7 to 4.3%) among females and slightly less, 3.4% per year among males (AAPC 95% CI 3.0 to 3.9%, p=0.02 for the difference between groups). **Figure 1e** demonstrates test use by IMD deprivation quintile. Test rates were highest in the most deprived cohort (quintile 5) and increased the most, with an AAPC of 4.4% (95% CI 4.1 to 4.8%), compared with those from the lower quintiles of deprivation (**Appendix Table 1**).

**Figure 1.**
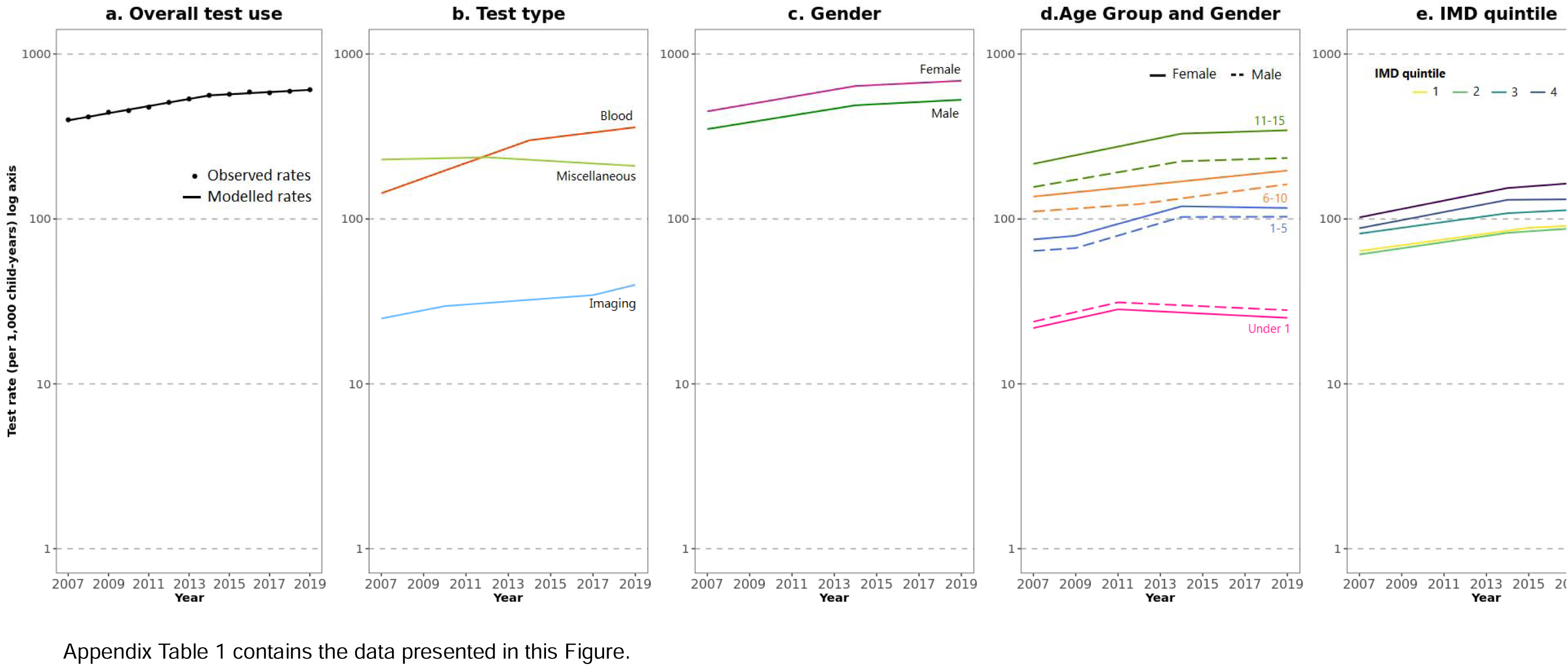
Temporal trends in paediatric test use in English primary care practices from 2007 to 2019.

### Temporal trends in specific tests

The average annual percentage change (AAPC) for each test is presented in Figure 2. The three greatest increases were for tests with zero or negligible use at the beginning of the study period. Fecal calprotectin testing was subject to the greatest average annual change, increasing from 0 tests/1,000 child-years in 2007 to 1.8 tests/1,000 child-years by 2019, equivalent to 105.5% per year (AAPC 95% CI 97.5 to 122.2%). This was followed by fractional exhaled nitric oxide (FeNO) tests which increased by 40.3% per year (AAPC 95% CI 26.7 to 64.7%) from 0 tests/1,000 child-years in 2007 to 0.2 tests/1,000 child-years in 2019, then vitamin D tests which increased by 27.0% per year (AAPC 95% CI 25.5 to 30.4%) from 0.5 tests/1,000 child-years in 2007 to 8.5 tests/1,000 child-years in 2019. The following tests increased by greater than 10% per year, in descending order: folate, vitamin B12, coeliac testing, helicobacter testing, iron studies, HbA1c, immunoglobulins, C reactive protein, MRI brain, bone profile, and allergen-specific IgE (see Appendix Table 2). Tests that decreased in use included urine microscopy/culture/sensitivities, hearing tests, spirometry, CT head, peak flow measurements, renal ultrasound, and monospot testing for glandular fever. The changes were largely consistent by gender, age group (see Appendix Figure 1), and deprivation quintile (see Appendix Figure 2).

**Figure 2.**
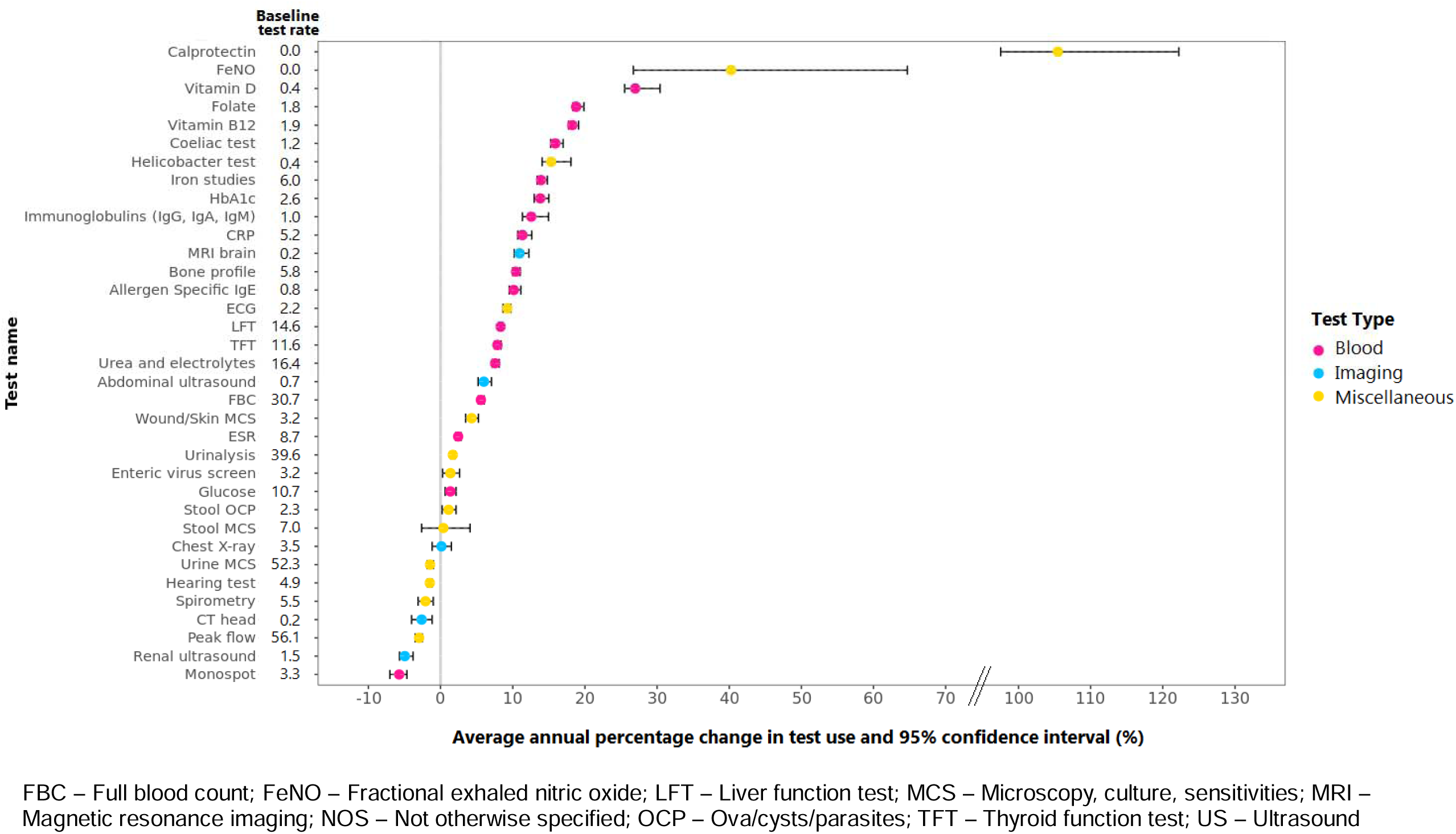
Temporal change in specific tests for children aged 0 to 15 years in English primary care from 2007 to 2019.

### Practice variation in test use

In 2019, 1,464 practices contributed 2,406,042 child-years of observation (ranging from 11 to 19,553 child-years per practice). The mean rate of test use by practice (adjusted for median age, gender, and deprivation level) was 609 tests per 1,000 child-years (standard deviation 41). Rates of testing varied from 0 to 2,249 tests per 1,000 child-years prior to adjustment, but after adjustment the range narrowed to 424 to 732 tests per 1,000 child-years (**Appendix Figure 3)**.

### Rank order of practice variability of specific tests

Figure 3 shows the rank order of the tests from highest to lowest practice variability (CoV). FeNO was subject to the greatest practice variability, with an adjusted CoV of 123.7% (95% CI 123.6 to 123.9%). This was followed by hearing tests (CoV 51.6%, 95% CI 51.4% to 51.7%) and Vitamin D tests (CoV 38.1%, 95% CI 38.0 to 38.3%). Tests with higher rates of use (represented by larger bubbles in Figure 3) were, on average, subject to lower practice variability.

**Figure 3.**
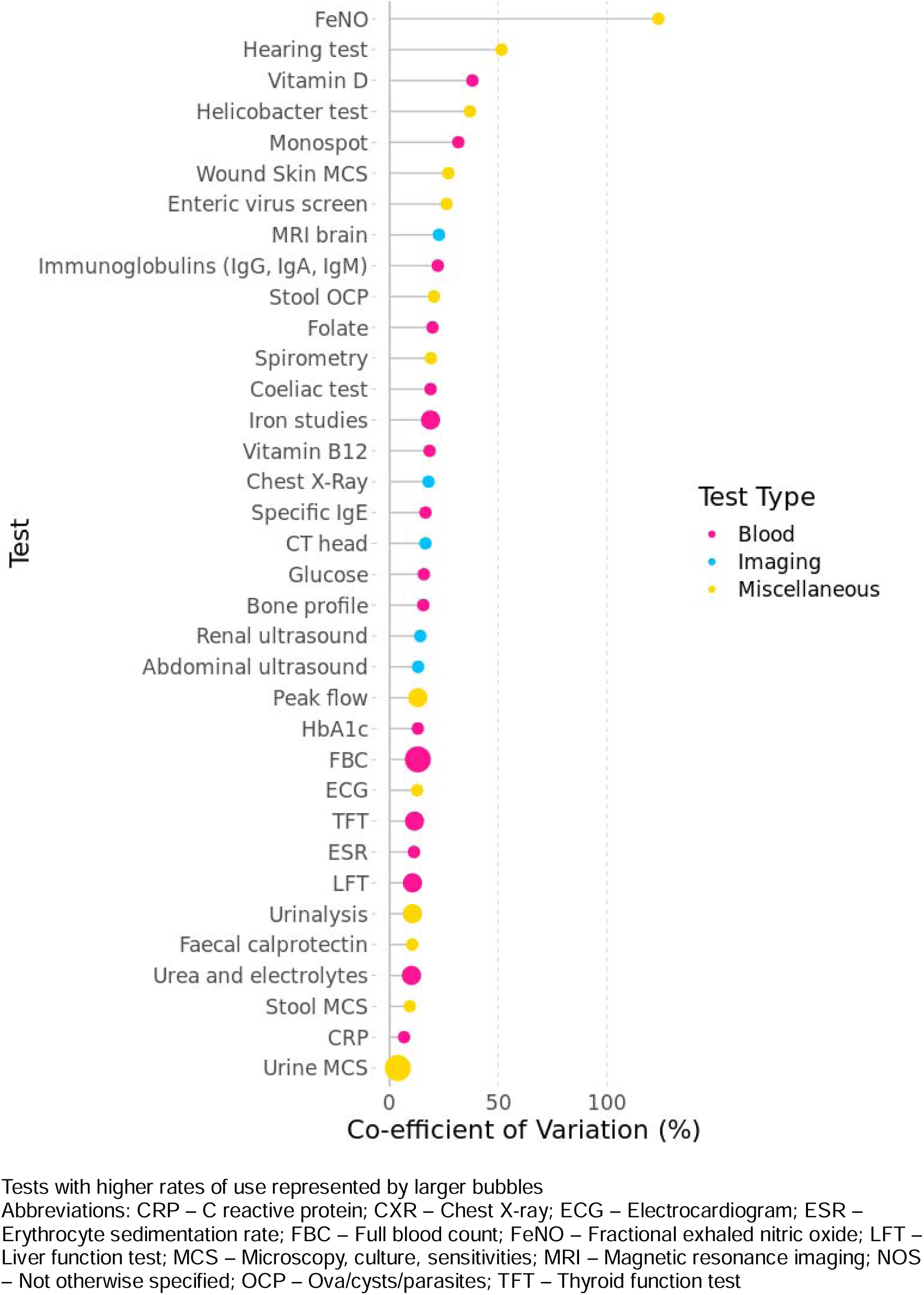
Rank order of between-practice variability of tests in 2019; adjusted for gender, age, and deprivation.

Figure 4 plots the adjusted coefficient of variation of each of the 35 tests against their test rate. The median coefficient of variation of test use was 16.5% (IQR 12.1 to 21.3%) and the median rate of test use was 6.9 tests/1000 child-years (IQR 2.4 to 19.3%). Most tests were either classified as low-test rate-high variability (37%, 13 out of 35) or high test rate-low variability (37%, 13 out of 35), see **Appendix Table 3**. The following five tests were classified as high test rate – high variability: Iron studies, Coeliac testing, Vitamin B_12_, Folate, and Vitamin D.

**Figure 4.**
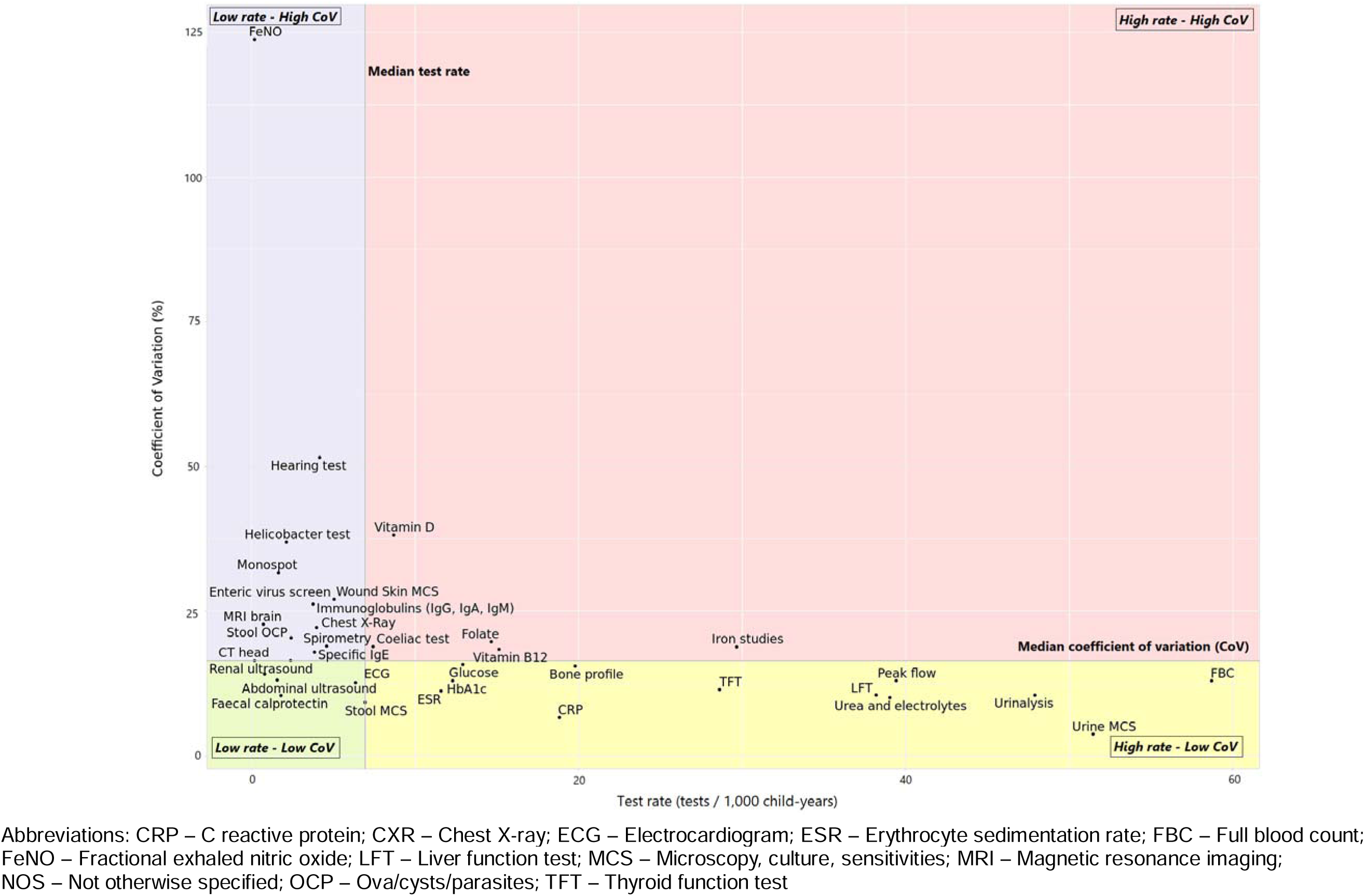
Test rate and degree of practice variability for specific tests in 2019.

## Discussion

To our knowledge, this study represents the first nationwide analysis of temporal trends and practice variation in paediatric test use between 2007 and 2019. We analysed 14 million tests over 27.8 million child-years of observation across nearly 1500 primary care practices and found that test use increased at a rate of 4% per year. Blood tests increased by the highest margin, and females aged 11-15 experienced the greatest increase, as well as children from practices in more deprived areas. Tests with the biggest temporal increases included: fecal calprotectin, FeNO, vitamin D, folate, vitamin B_12_, coeliac, and helicobacter tests. Tests subject to the largest practice variation included FeNO, hearing tests, vitamin D, helicobacter testing, and monospot testing for glandular fever. We also identified tests with high rates of test use and practice variability: iron studies, vitamin B_12_, coeliac test, folate, and vitamin D.

The increasing use of both calprotectin and FeNO tests reflect the implementation of these tests in UK primary care during the study period. These two tests serve as interesting case studies: in 2019 there was low practice variability in calprotectin testing but high variability in FeNO testing. Several factors may explain this discrepancy in practice variability in 2019, including the ease with which both test samples are obtained and analysed. Additionally, it could be due to the lack of equipment, time, access, and funding for FeNO testing in primary care despite evidence of their feasibility and acceptability, and national guidance recommending FeNO testing for the diagnostic workup of childhood asthma. [23–25]

The rise in vitamin D tests is consistent with the 42-fold increase observed for children in Minnesota from 2002 to 2017, though the odds of detecting low levels remained small, suggesting that tests were overused.[26] Similar trends were observed in Australia and Canada.[27, 28] There are no specific guidelines for the paediatric population, however, the US Preventive Services Task Force and the American Society for Clinical Pathology guidelines for adults do not recommend screening for vitamin D deficiency [29]. Therefore, increased vitamin D testing is likely to represent overuse in children.

The increases observed in requests for hematinic tests (vitamin B_12_, folate, iron) were surprising, and indicate growing clinician concern for nutritional deficiencies and anaemia. Similar test increases were observed in adults in UK primary care [11] which could suggest a creep of adult diagnostic practices into paediatric care.[18] Haematinic tests, in addition to testing for coeliac disease, helicobacter pylori, vitamin D, CRP, and HbA1c may also be requested for more non-specific symptoms including fatigue, musculoskeletal issues or abdominal symptoms. Further research is needed to understand the implications of the observed increases in testing and examine their appropriateness. This could be achieved by examining the test results and disease sequelae (similar to the Vitamin D studies described earlier[26, 28]) or determining whether testing indications were concordant with evidence-based guidelines.

Previous research on paediatric diagnostic test use focused on specific tests, were undertaken in smaller geographic areas, or in hospital settings. Quality improvement studies were conducted in paediatric intensive care units to reduce blood cultures in febrile infants; which drive unnecessary antibiotic treatment and hospital-acquired infections.[30] Potassium testing was also examined in a retrospective study which revealed that increased potassium testing did not influence the need for potassium replacement in patients with status asthmaticus on continuous albuterol.[31] Increased use of certain tests can be appropriate in the right population and setting. For example, rapid diagnostic testing for respiratory infections in the emergency setting has been shown to prevent unnecessary antibiotic prescriptions. [32] It is, therefore, prudent to examine all test use to identify potential areas where testing may confer benefit as well as areas where testing causes harms.

Our study results concur with findings of increased blood test utilisation in Oxfordshire primary care practices; particularly for vitamin D, folate, vitamin B_12_, iron studies, coeliac testing, HbA1c, bone profile, CRP, thyroid function tests, urea and electrolytes, and liver function tests.[12] Notably, test increases were more pronounced in our nationwide study compared with Oxfordshire. This may be the result of local policies discouraging testing in primary care or the referral of children to outpatient services for testing.

We specifically looked at test utilisation in the period preceding the pandemic, however, testing rates likely decreased substantially during the pandemic with decreased paediatric consultations in primary care.[33] Assuming test rates recovered, and test rates continued at the rate of growth since 2014 (APC 1.6%), then by 2024 test rates in primary care would be 656 tests/1,000 children per year. This has considerable cost implications. A Vitamin D test costs approximately £10 at the Oxford University Hospitals NHS Trust Laboratories. Applying the rates of vitamin D requests to the rest of the UK population in 2019, vitamin D tests requested for children in primary care cost £2.2 million in 2019. If rates continued to increase at the AAPC of 29.6% per year, then by 2024, the rate of vitamin D testing would be 42.3 tests/1,000 child-years, costing £5.4 million across UK primary care annually. While these are estimates (and assume testing rates recovered post-pandemic), they provide an indication of the potential scale of the financial consequences if testing rates continued to increase. Additional harms, including physical side effects of tests from radiation, psychological distress and anxiety associated with tests, and the potential consequences that include overdiagnosis, further tests, and unnecessary treatments are also important and merit further research.

This study had several limitations. First, CPRD data relies on the quality of the electronic data input by clinicians using electronic health record software which can be highly variable. Second, double counting could have occurred if both the request and the completed test were coded separately, however, this is not likely to have varied by patient demographic, practice, or calendar time, therefore, the metrics of temporal trends and practice variation would nevertheless be valid. Third, there was some subjectivity in how the test names were coded and grouped. To address this, the code list of tests (and their associated panels) was developed and cross-checked with 1) existing NHS trust laboratory test lists, 2) the test codes from a previous laboratory-based study based in an NHS trust corroborated by a consultant chemical pathologist[12] and 3) another clinician-researcher (CH). Grouped test codes (i.e. “Immunoglobulins (IgA, IgG, IgM)”) may have obscured more subtle trends in contrast with single condition-specific test codes, which can reveal more about the diagnostic strategies for a particular disease. For example, the decline in monospot testing for glandular fever may reflect shifts in clinicians’ preferences towards alternative diagnostic strategies such as serology tests or clinical diagnosis.

## Conclusions

This study provides a broader picture of paediatric testing practices based on individual-level data across primary care in England. Increased testing rates can generate more clinical activity including more specialist referrals, and the potential cost implications are substantial. Future research should compare tests against clinical guideline standards and examine the test results to judge whether test increases are warranted and evaluate its downstream impact on patient outcomes and cost.

## Study approval

The protocol was approved via the CPRD’s Research Data Governance process (study reference 22_001998) and is available on the Open Science Framework (osf.io/pwgtf).

## Data Availability

The study protocol was registered on the Open Science Framework (doi:10.17605/osf.io/gkncj). All test codes, R code used for data management, analysis and creating the figures is available on GitHub (https://github.com/elizabethtthomas/cprd-paediatric-tests). Applications for the original dataset is subject to approval by CPRD.

https://doi.org/10.17605/osf.io/gkncj

## Data sharing and availability

The study protocol was registered on the Open Science Framework (doi:10.17605/osf.io/gkncj). All test codes, R code used for data management, analysis and creating the figures is available on GitHub (https://github.com/elizabethtthomas/cprd-paediatric-tests).

## Acknowledgements

The authors would like to thank Dr Cynthia Wright-Drakesmith for her assistance with data extraction and cleaning.

## Author contributions

ETT contributed to study conceptualisation, methodology, completed the data analyses and wrote the original draft. DRW reviewed the statistical aspects of the study and provided critical input on the original manuscript. PG provided supervisory input and comments and feedback on the manuscript. RP provided supervisory input, contributed to conceptualisation, methodology, reviewed the statistical aspects of the study and revision of the manuscript. CH also provided supervisory input, and contributed to conceptualisation, methodology, data interpretation and revision of the manuscript. All authors had full access to all the data in the study and had final responsibility for the decision to submit for publication.

## Declarations of interests

This project was funded by a grant from the NIHR SPCR grant (Award 624). The funders were not involved in the study design and conduct, data collection and interpretation, or manuscript preparation and approval to submit the manuscript for publication.

ETT was supported by a Clarendon scholarship to study for a Doctor of Philosophy (DPhil) at the University of Oxford (2020-23). PG has received grants from the Canadian Institutes of Health Research (CIHR), the Physicians Services Incorporated Foundation, and The Hospital for Sick Children. He has received nonfinancial support from the EBMLive Steering Committee (expenses reimbursed to attend conferences) and the CIHR Institute of Human Development, Child and Youth Health (as a member of the institute advisory board, expenses reimbursed to attend meetings), is a member of the CMAJ Open and BMJ Evidence Based Medicine Editorial Board. RP is partly supported by the NIHR Applied Research Collaboration (ARC) Oxford & Thames Valley, the NIHR Oxford BRC, the NIHR Oxford MedTech and In-Vitro Diagnostics Co-operative (MIC) and the Oxford Martin School. CJH receives funding support from the NIHR School of Primary Care Research. The funders had no role in study design, manuscript submission, or collection, management, analysis, or interpretation of study data. All other authors have no sources of funding to declare.

## Notes

### Clinical Protocols

https://github.com/elizabethtthomas/cprd-paediatric-tests

### Author Declarations

The protocol was approved via the Clinical Practice Research Datalink Research Data Governance process (study reference 22_001998).

## References

1. UK Department of Health. Report of the Review of NHS Pathology Services in England chaired by Lord Carter of Coles. London; 2006.

2. Geist R, Weinstein M, Walker L, Campo J V. Medically unexplained symptoms in young people: The doctor’s dilemma. Paediatr Child Health. 2008;13.

3. Foot C, Naylor C, Imison C. The quality of GP diagnosis and referral. London, UK: The King’s Fund. 2010.

4. Hiscock H, Perera P, McLean K, Roberts G. Evidence Check: Variation in paediatric clinical practice. 2015.

5. Hiscock H, Perera P, Mclean K, Roberts G, Lucas G, Kelly M, et al. Variation in paediatric clinical practice: A review of care in inpatient, outpatient and emergency department settings. Journal of Paediatrics and Child Health. 2016;52:691–3.

6. Public Health England. The 2nd Atlas of Variation in NHS Diagnostic Services in England. NHS RightCare. 2017; January.

7. Fradet C, McGrath PJ, Kay J, Adams S, Luke B. A prospective survey of reactions to blood tests by children and adolescents. Pain. 1990;40.

8. Ralston SL, Schroeder AR. Why it is so hard to talk about overuse in pediatrics and why it matters. JAMA Pediatrics. 2017;171.

9. Grant AM, Wright FA, Chapman LRM, Cook E, Byrne R, O’Brien TA. Rethinking Blood Testing in Pediatric Cancer Patients: A Quality Improvement Approach. Pediatr Qual Saf. 2022;7.

10. O’Sullivan JW, Stevens S, Hobbs FDR, Salisbury C, Little P, Goldacre B, et al. Temporal trends in use of tests in UK primary care, 2000-15: Retrospective analysis of 250 million tests. The BMJ. 2018;363.

11. O’Sullivan JW, Stevens S, Oke J, Hobbs FDR, Salisbury C, Little P, et al. Practice variation in the use of tests in UK primary care: A retrospective analysis of 16 million tests performed over 3.3 million patient years in 2015/16. BMC Med. 2018;16.

12. Thomas ET, Withrow DR, Shine B, Gill P, Perera R, Heneghan C. Trends in diagnostic tests ordered for children: a retrospective analysis of 1.7 million laboratory test requests in Oxfordshire, UK from 2005 to 2019. Arch Dis Child. 2023;archdischild-2023-325550.

13. Clinical Practice Research Datalink. CPRD Aurum February 2022 (Version 2022.02.001). CPRD. 2022. 10.48329/gcgx-f815. Accessed 16 Dec 2023.

14. Ruzangi J, Blair M, Cecil E, Greenfield G, Bottle A, Hargreaves DS, et al. Trends in healthcare use in children aged less than 15 years: A population-based cohort study in England from 2007 to 2017. BMJ Open. 2020;10.

15. Wolf A, Dedman D, Campbell J, Booth H, Lunn D, Chapman J, et al. Data resource profile: Clinical Practice Research Datalink (CPRD) Aurum. Int J Epidemiol. 2019;48.

16. Clinical Practice Research Datalink. Small area level data based on practice postcode: Documentation and Data Dictionary. https://cprd.com/sites/default/files/2022-05/Documentation_SmallAreaData_Practice_set22_v3.4_1.pdf.2022.

17. Herrett E, Gallagher AM, Bhaskaran K, Forbes H, Mathur R, Staa T van, et al. Data Resource Profile: Clinical Practice Research Datalink (CPRD). Int J Epidemiol. 2015;44.

18. Thomas ET, Glogowska M, Hayward G, Gill P, Perera R, Heneghan C. General practitioners’ perspectives on diagnostic tests for children: a qualitative interview study. British Journal of General Practice. 2024;In Press.

19. Thomas ET, Thomas ST, Perera R, Gill PJ, Moloney S, Heneghan C. The quality of diagnostic guidelines for children in primary care: A meta-epidemiological study. J Paediatr Child Health. 2023;59:1053–60.

20. Thomas ET, Thomas ST, Perera R, Gill PJ, Moloney S, Heneghan CJ. The quality of paediatric asthma guidelines: evidence underpinning diagnostic test recommendations from a meta-epidemiological study. Fam Pract. 2023. 10.1093/fampra/cmad052.

21. National Cancer Institute, Sciences D of CC and P. Joinpoint Regression Program. USA: Surveillance Research Program. 2020.

22. R Core Team. R software. 2021.

23. Lo D, Beardsmore C, Roland D, Richardson M, Yang Y, Danvers L, et al. Spirometry and FeNO testing for asthma in children in UK primary care: A prospective observational cohort study of feasibility and acceptability. British Journal of General Practice. 2020;70.

24. NICE. Overview | Asthma: diagnosis, monitoring and chronic asthma management | Guidance | NICE. National Institute for Health and Care Excellence. 2017; November.

25. Petsky HL, Kew KM, Chang AB. Exhaled nitric oxide levels to guide treatment for children with asthma. Cochrane Database of Systematic Reviews. 2016;2016.

26. Kerber AA, Pitlick MM, Kellund AE, Weaver AL, Kumar S, Joshi AY. Stable Rates of Low Vitamin D Status Among Children Despite Increased Testing: A Population-Based Study. Journal of Pediatrics. 2021;239.

27. Canadian Health Measures Survey. Canadian Health Measures Survey: Vitamin D status of Canadians, 2007 to 2019. Statistics Canada,. 2023. https://www150.statcan.gc.ca/n1/daily-quotidien/231128/dq231128e-eng.htm. Accessed 30 Jan 2024.

28. Zurynski Y, Munns CF, Sezgin G, Imai C, Georgiou A. Vitamin D testing in children and adolescents in Victoria, Australia: are testing practices in line with global recommendations? Arch Dis Child. 2023;:archdischild-2022-325000.

29. Krist AH, Davidson KW, Mangione CM, Cabana M, Caughey AB, Davis EM, et al. Screening for Vitamin D Deficiency in Adults: US Preventive Services Task Force Recommendation Statement. JAMA - Journal of the American Medical Association. 2021;325.

30. Woods-Hill CZ, Colantuoni EA, Koontz DW, Voskertchian A, Xie A, Thurm C, et al. Association of Diagnostic Stewardship for Blood Cultures in Critically Ill Children with Culture Rates, Antibiotic Use, and Patient Outcomes: Results of the Bright STAR Collaborative. JAMA Pediatr. 2022;176.

31. Cox C, Patel K, Cantu R, Akmyradov C, Irby K. Hypokalemia Measurement and Management in Patients With Status Asthmaticus on Continuous Albuterol. Hosp Pediatr. 2022;12.

32. Weragama K, Mudgil P, Whitehall J. Diagnostic Stewardship—The Impact of Rapid Diagnostic Testing for Paediatric Respiratory Presentations in the Emergency Setting: A Systematic Review. Children. 2022;9.

33. Foley KA, Maile EJ, Bottle A, Neale FK, Viner RM, Kenny SE, et al. Impact of COVID-19 on primary care contacts with children and young people in England: longitudinal trends study 2015-2020. British Journal of General Practice. 2022;72.

