## Appendices for "Temporal trends and practice variation of paediatric diagnostic tests in primary care"

**Appendix 1 Sources of included tests**

| **Rank** | **Test name** | **Number of tests** | |  | **Source** |
| --- | --- | --- | --- | --- | --- |
| 1 | Urine MCS | | 1391785 |  | Top 25* |
| 2 | Urinalysis | | 1261359 |  |  |
| 3 | Full blood count | | 1249322 |  |  |
| 4 | Peak flow | | 1184109 |  |  |
| 5 | Urea and electrolytes | | 790364 |  |  |
| 6 | Liver function test | | 760041 |  |  |
| 8 | Thyroid function tests | | 560244 |  |  |
| 9 | Iron studies | | 489989 |  |  |
| 10 | Bone profile | | 375914 |  |  |
| 11 | Glucose | | 361960 |  |  |
| 12 | C reactive protein | | 349276 |  |  |
| 13 | Erythrocyte sedimentation rate | | 314392 |  |  |
| 14 | Vitamin B_12_ | | 217619 |  |  |
| 15 | Stool MCS | | 214476 |  |  |
| 16 | Folate | | 204739 |  |  |
| 17 | Vitamin D | | 168323 |  |  |
| 18 | HbA1c | | 162022 |  |  |
| 19 | Wound/Skin MCS | | 138975 |  |  |
| 20 | Spirometry | | 131770 |  |  |
| 21 | Hearing test | | 130290 |  |  |
| 22 | Coeliac test | | 120337 |  |  |
| 23 | Enteric virus screen | | 117247 |  |  |
| 25 | ECG | | 105247 |  |  |
| 26 | Chest X-Ray | | 104604 |  |  |
| 29 | Stool OCP | | 92839 |  |  |
| 33 | Immunoglobulins (IgG, IgA, IgM) | | 70549 |  | Interview |
| 34 | Monospot | | 68121 |  | Literature |
| 41 | Allergen Specific IgE | | 42642 |  | Interview |
| 49 | Abdominal ultrasound | | 34195 |  | Literature |
| 54 | Helicobacter test | | 30655 |  | Interview |
| 57 | Renal ultrasound | | 27606 |  | Literature |
| 80 | Calprotectin | | 14637 |  | Interview |
| 84 | MRI head | | 12936 |  | Literature |
| 115 | CT head | | 5437 |  | Literature |
| 197 | Fractional exhaled nitric oxide | | 977 |  | Interview |

*Excludes unspecified tests

| Appendix Table 1 Standardised rates of test use overall; stratified by test type, gender, age, and deprivation quintile; annual percentage change and average annual percentage change from 2007 to 2019 | | | | | | | | |
| --- | --- | --- | --- | --- | --- | --- | --- | --- |
|  |  |  | **Rate* (per 1,000 child years)** | |  | **95% Confidence Interval** | |  |
|  | **Start** | **End** | **Start** | **End** | **APC (%)** | **Lower Limit** | **Upper Limit** | **AAPC (%) and 95% CI** |
| Overall | 2007 | 2014 | 399.2 | 561.9 | 5.1 | 4.7 | 5.6 | 3.6 (3.4, 3.8) |
|  | 2014 | 2019 | 561.9 | 607.7 | 1.6 | 0.9 | 2.1 |  |
| Blood Tests | 2007 | 2014 | 144,6 | 296.7 | 11.1 | 10.3 | 12 | 8.0 (7.7, 8.4) |
|  | 2014 | 2019 | 296.7 | 356.9 | 3.7 | 2.8 | 4.6 |  |
| Imaging | 2007 | 2010 | 25.3 | 29.8 | 5.9 | 4.3 | 8.8 | 4.0 (3.7, 4.3) |
|  | 2010 | 2017 | 29.8 | 34.4 | 2.2 | 1.4 | 2.6 |  |
|  | 2017 | 2019 | 34.4 | 39.9 | 7.4 | 5.2 | 9.1 |  |
| Miscellaneous | 2007 | 2012 | 229.3 | 236.2 | 0.6 | -0.2 | 2.8 | -0.7 (-1.1, -0.3) |
|  | 2012 | 2019 | 236.2 | 210.8 | -1.7 | -2.7 | -1.2 |  |
| Female | 2007 | 2014 | 448.6 | 638.7 | 5.2 | 4.7 | 5.7 | 3.6 (3.4, 3.9) |
|  | 2014 | 2019 | 638.7 | 690.6 | 1.5 | 0.8 | 2.1 |  |
| Male | 2007 | 2014 | 356.3 | 491.2 | 4.9 | 4.4 | 5.4 | 3.5 (3.2, 3.7) |
|  | 2014 | 2019 | 491.2 | 528.4 | 1.5 | 0.8 | 2.2 |  |
| Female |  |  |  |  |  |  |  |  |
| <1 | 2007 | 2011 | 21.5 | 27.3 | 6.8 | 4.4 | 10.3 | 1.2 (0.6, 1.8) |
|  | 2011 | 2019 | 27.3 | 25.5 | -1.5 | -2.5 | -0.6 |  |
| 1-5 | 2007 | 2009 | 75.6 | 78.8 | 2.7 | -1.0 | 7.5 | 3.7 (3.0, 4.4) |
|  | 2009 | 2014 | 78.8 | 117.9 | 8.6 | 6.5 | 10.0 |  |
|  | 2014 | 2019 | 117.9 | 118.6 | -0.5 | -2.9 | 0.8 |  |
| 6-10 | 2007 | 2019 | 136.3 | 196.4 | 3.1 | 2.3 | 3.9 | 3.1 (2.3, 3.9) |
| 11-15 | 2007 | 2014 | 215.2 | 328.6 | 6.2 | 5.6 | 6.8 | 4.0 (3.7, 4.3) |
|  | 2014 | 2019 | 328.6 | 350.1 | 0.9 | -0.1 | 1.9 |  |
| Male |  |  |  |  |  |  |  |  |
| <1 | 2007 | 2011 | 23.5 | 30.0 | 7.0 | 4.3 | 11.5 | 1.3 (0.7, 2.1) |
|  | 2011 | 2019 | 30.0 | 28.7 | -1.4 | -2.6 | -0.4 |  |
| 1-5 | 2007 | 2009 | 64.8 | 66.9 | 2.0 | -1.4 | 5.9 | 4.1 (3.4, 4.6) |
|  | 2009 | 2014 | 66.9 | 101.8 | 9.1 | 7.4 | 10.3 |  |
|  | 2014 | 2019 | 101.8 | 105.2 | 0.1 | -2.1 | 1.3 |  |
| 6-10 | 2007 | 2012 | 108.6 | 121.0 | 2.1 | -0.8 | 3.3 | 3.2 (2.8, 3.7) |
|  | 2012 | 2019 | 121.0 | 159.1 | 4.0 | 3.3 | 6.7 |  |
| 11-15 | 2007 | 2014 | 159.5 | 226.7 | 5.3 | 4.5 | 6.4 | 3.4 (3.0, 3.9) |
|  | 2014 | 2019 | 226.7 | 235.5 | 0.9 | -0.9 | 2.1 |  |
| IMD 1 (least deprived quintile) | 2007 | 2015 | 63.5 | 87.9 | 4.1 | 3.7 | 4.7 | 3.2 (3.0, 3.5) |
|  | 2015 | 2019 | 87.9 | 94.6 | 1.5 | 0.1 | 2.4 |  |
| IMD 2 | 2007 | 2014 | 61.4 | 83.5 | 4.4 | 3.9 | 5.6 | 3.4 (3.1, 3.8) |
|  | 2014 | 2019 | 83.5 | 92.4 | 2.0 | 0.5 | 2.8 |  |
| IMD 3 | 2007 | 2014 | 82.4 | 109.0 | 4.2 | 3.7 | 5.0 | 3.0 (2.8, 3.4) |
|  | 2014 | 2019 | 109.0 | 116.7 | 1.5 | 0.4 | 2.2 |  |
| IMD 4 | 2007 | 2014 | 88.5 | 130.1 | 5.8 | 5.1 | 6.7 | 3.5 (3.1, 3.9) |
|  | 2014 | 2019 | 130.1 | 134.4 | 0.3 | -0.9 | 1.3 |  |
| IMD 5 (most deprived quintile) | 2007 | 2014 | 103.4 | 154.4 | 6.0 | 5.4 | 7.0 | 4.4 (4.1, 4.8) |
|  | 2014 | 2019 | 154.4 | 169.6 | 2.2 | 0.9 | 3.1 |  |

*Observed rates; Abbreviations: APC – annual percentage change; AAPC – average annual percentage change

Appendix Table 2 Average annual percentage change and test rates of 35 specific tests for children aged 0 to 15 years in primary care from 2007 to 2019

| **Test name** | **Test type** | **AAPC** | **95% CI lower limit** | **95% CI upper limit** | **2007 test rate* (tests/1,000 child years)** | **2019 test rate* (tests/1,000 child years)** |
| --- | --- | --- | --- | --- | --- | --- |
| Calprotectin | Miscellaneous | 105.5 | 97.5 | 122.2 | 0ª | 1.8 |
| FeNO | Miscellaneous | 40.3 | 26.7 | 64.7 | 0ª | 0.2 |
| Vitamin D | Blood | 27.0 | 25.5 | 30.4 | 0.4 | 8.5 |
| Folate | Blood | 18.8 | 18.4 | 19.9 | 1.8 | 14.5 |
| Vitamin B_12_ | Blood | 18.3 | 17.7 | 19.1 | 1.9 | 15 |
| Coeliac test | Blood | 15.9 | 15.3 | 17.0 | 1.2 | 7.5 |
| Helicobacter test | Miscellaneous | 15.3 | 14.1 | 18.1 | 0.4 | 2.1 |
| Iron studies | Blood | 13.9 | 13.4 | 14.8 | 6 | 29.5 |
| HbA1c | Blood | 13.8 | 13.0 | 15.0 | 2.6 | 12.3 |
| Immunoglobulins (IgG, IgA, IgM) | Blood | 12.6 | 11.3 | 15.0 | 1 | 4 |
| C reactive protein | Blood | 11.3 | 10.7 | 12.6 | 5.2 | 18.8 |
| MRI brain | Imaging | 11.0 | 10.2 | 12.2 | 0.2 | 0.7 |
| Bone profile | Blood | 10.5 | 10.0 | 11.0 | 5.8 | 19.7 |
| Allergen Specific IgE | Blood | 10.1 | 9.5 | 11.1 | 0.8 | 2.37 |
| Electrocardiogram | Miscellaneous | 9.2 | 8.7 | 9.7 | 2.2 | 6.4 |
| Liver function test | Blood | 8.3 | 8.1 | 8.6 | 14.6 | 38.1 |
| Thyroid function test | Blood | 7.9 | 7.5 | 8.4 | 11.6 | 28.5 |
| Urea and electrolytes | Blood | 7.6 | 7.2 | 8.1 | 16.4 | 38.9 |
| Abdominal ultrasound | Imaging | 6.0 | 5.2 | 7.1 | 0.7 | 1.56 |
| Full blood count | Blood | 5.6 | 5.2 | 6.0 | 30.7 | 58.5 |
| Wound/Skin MCS | Miscellaneous | 4.3 | 3.5 | 5.2 | 3.2 | 5.1 |
| Erythrocyte sedimentation rate | Blood | 2.4 | 2.1 | 2.8 | 8.7 | 11.6 |
| Urinalysis | Miscellaneous | 1.7 | 1.4 | 2.0 | 39.6 | 48 |
| Enteric virus screen | Miscellaneous | 1.4 | 0.3 | 2.6 | 3.2 | 3.8 |
| Glucose | Blood | 1.4 | 0.6 | 2.1 | 10.7 | 12.8 |
| Stool OCP | Miscellaneous | 1.1 | 0.2 | 2.1 | 2.3 | 2.4 |
| Stool MCS | Miscellaneous | 0.4 | -2.6 | 4.1 | 7 | 6.9 |
| Chest X-ray | Imaging | 0.1 | -1.2 | 1.5 | 3.5 | 3.8 |
| Urine MCS | Miscellaneous | -1.5 | -1.9 | -1.0 | 52.3 | 44.9 |
| Hearing test | Miscellaneous | -1.5 | -1.8 | -1.2 | 4.9 | 4.1 |
| Spirometry | Miscellaneous | -2.1 | -3.1 | -1.0 | 5.5 | 4.6 |
| CT head | Imaging | -2.6 | -4.0 | -1.2 | 0.2 | 0.2 |
| Peak flow | Miscellaneous | -3.0 | -3.5 | -2.5 | 56.1 | 39.4 |
| Renal ultrasound | Imaging | -5.0 | -5.7 | -3.8 | 1.5 | 0.8 |
| Monospot | Blood | -5.8 | -7.0 | -4.7 | 3.3 | 1.6 |
| *Observed rates; Abbreviations: AAPC – average annual percentage change; CT – computed tomography; FeNO – fractional exhaled nitric oxide test for asthma; MCS – microscopy, culture, and sensitivities; MRI – magnetic resonance imaging; OCP – ova, cysts, and parasites | | | | | | |
| ª There were zero tests in 2007, the first recorded calprotectin test was in 2009 and the first recorded FeNO test was in 2010 | | | | | |  |

Appendix Figure 1 Temporal changes in specific tests for children aged 0 to 15 from 2007 to 2019; stratified by gender and age.

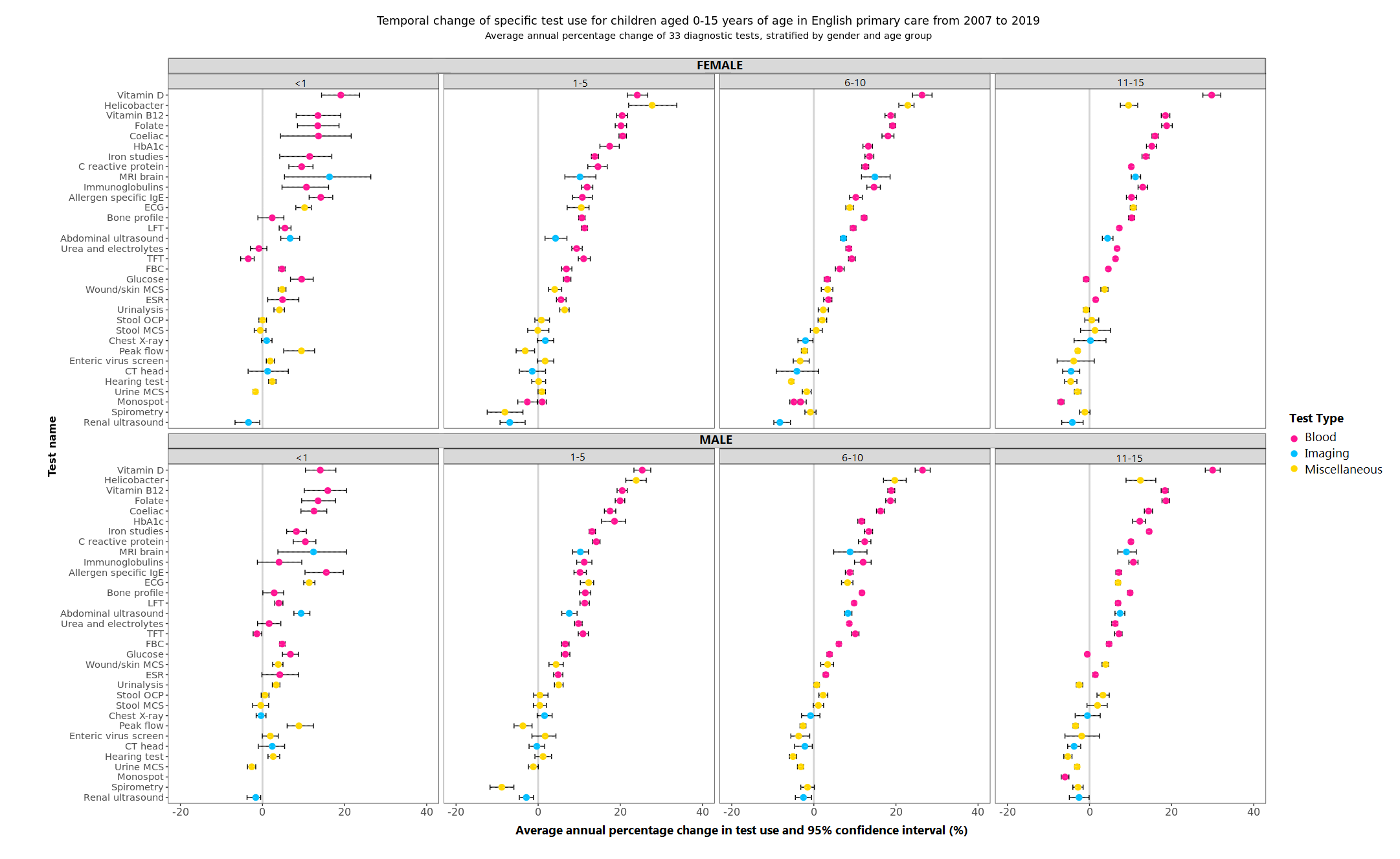

CRP – C reactive protein; ECG – Electrocardiogram; ESR – Erythrocyte sedimentation rate; FBC – Full blood count; LFT – Liver function test;

MCS – Microscopy, culture, sensitivities; NOS – Not otherwise specified; OCP – Ova/cysts/parasites; TFT – Thyroid function test; US – Ultrasound.

Tests were excluded if there were too few tests to perform a meaningful analysis, or the test was not technically possible to perform, leaving the outcome blank in the graph (e.g., spirometry in <1 year old).

Appendix Figure 2 Temporal changes in specific tests for children aged 0 to 15 from 2007 to 2019; stratified by Index of Multiple Deprivation Quintile.

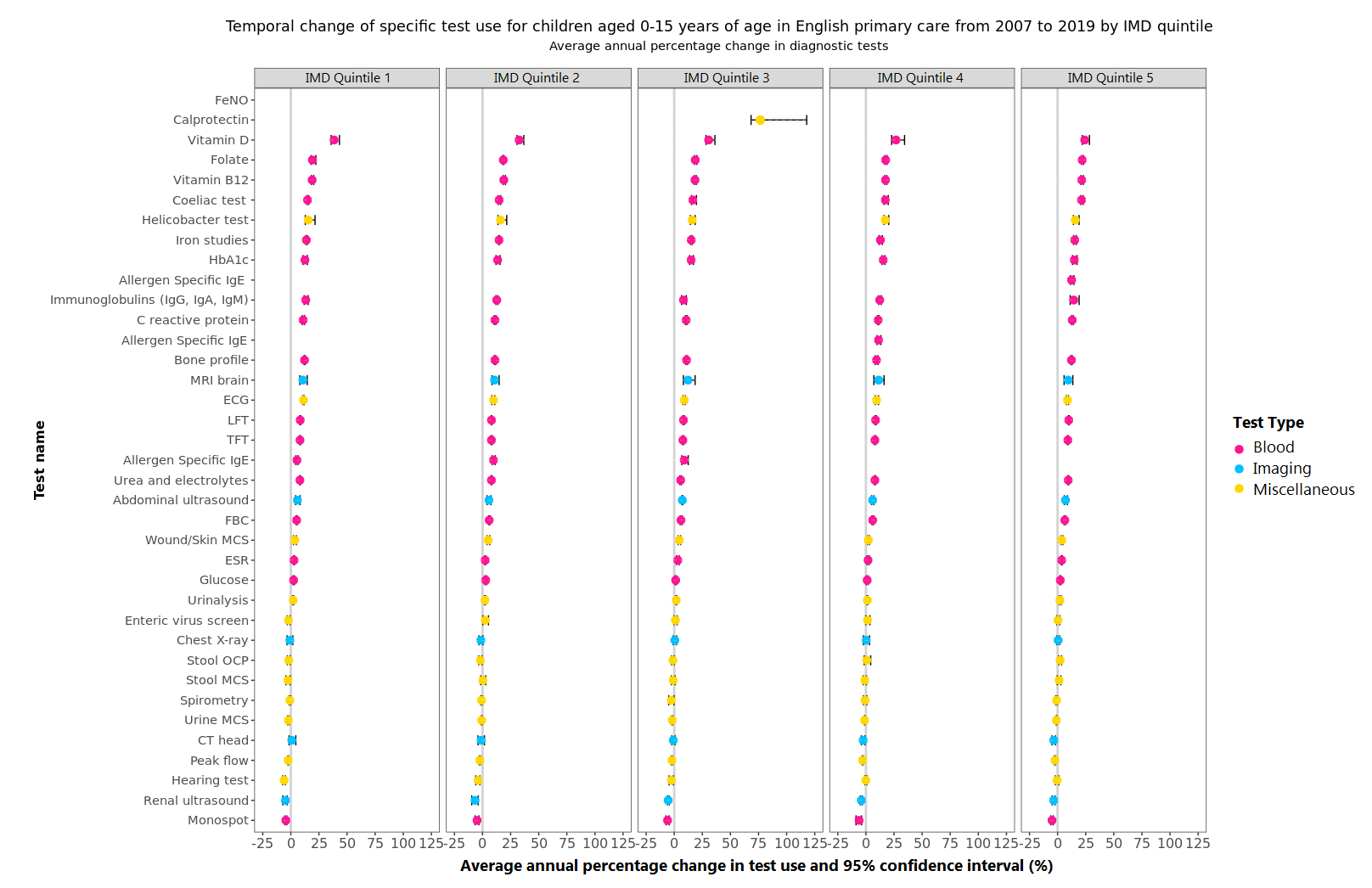

CRP – C reactive protein; ECG – Electrocardiogram; ESR – Erythrocyte sedimentation rate; FBC – Full blood count; FeNO – Fractional exhaled nitric oxide; LFT – Liver function test; MCS – Microscopy, culture, sensitivities; NOS – Not otherwise specified; OCP – Ova/cysts/parasites; TFT – Thyroid function test; US – Ultrasound.

Tests were excluded if there were too few tests to perform a meaningful analysis, leaving the outcome blank in the graph.

Appendix Figure 3 Crude and adjusted practice-specific test request rates for children aged 0 to 15 in 2019; Adjusted for gender, age, and deprivation

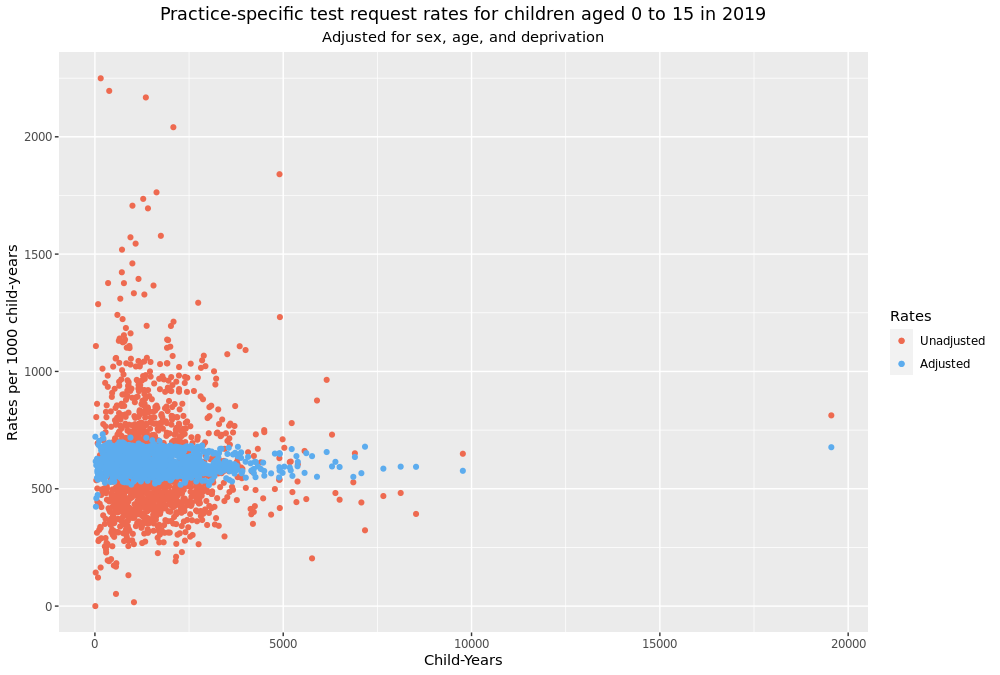

| Appendix Table 3 Adjusted test rates and coefficient of variation (CoV) with corresponding 95% confidence intervals in 2019 | | | | | | |
| --- | --- | --- | --- | --- | --- | --- |
| **Test name** | **Test type** | **Adjusted mean rate (tests/1,000 child-years)** | **Adjusted CoV (%)** | **95% CI lower limit (%)** | **95% CI upper limit (%)** | **Rate-Variability*** |
| FBC | Blood | 58.7 | 13.0 | 12.9 | 13.1 | High test rate - Low CoV |
| Urine MCS | Miscellaneous | 48.4 | 3.8 | 3.7 | 3.9 | High test rate - Low CoV |
| Urinalysis | Miscellaneous | 47.9 | 10.5 | 10.3 | 10.7 | High test rate - Low CoV |
| Peak flow | Miscellaneous | 39.4 | 13.0 | 12.8 | 13.3 | High test rate - Low CoV |
| Urea and electrolytes | Blood | 39.0 | 10.1 | 10.0 | 10.2 | High test rate - Low CoV |
| LFT | Blood | 38.2 | 10.5 | 10.4 | 10.7 | High test rate - Low CoV |
| Iron studies | Blood | 29.7 | 18.8 | 18.7 | 19.0 | High test rate - High CoV |
| TFT | Blood | 28.6 | 11.5 | 11.3 | 11.6 | High test rate - Low CoV |
| Bone profile | Blood | 19.8 | 15.5 | 15.4 | 15.7 | High test rate - Low CoV |
| CRP | Blood | 18.8 | 6.7 | 6.5 | 6.8 | High test rate - Low CoV |
| Vitamin B12 | Blood | 15.1 | 18.4 | 18.3 | 18.6 | High test rate - High CoV |
| Folate | Blood | 14.6 | 19.7 | 19.6 | 19.9 | High test rate - High CoV |
| Glucose | Blood | 12.9 | 15.8 | 15.7 | 15.9 | High test rate - Low CoV |
| HbA1c | Blood | 12.3 | 13.0 | 12.9 | 13.1 | High test rate - Low CoV |
| ESR | Blood | 11.6 | 11.2 | 11.1 | 11.4 | High test rate - Low CoV |
| Vitamin D | Blood | 8.7 | 38.1 | 38.0 | 38.3 | High test rate - High CoV |
| Coeliac | Blood | 7.4 | 18.9 | 18.8 | 19.0 | High test rate - High CoV |
| Stool MCS | Miscellaneous | 6.9 | 9.3 | 9.1 | 9.4 | High test rate - Low CoV |
| ECG | Miscellaneous | 6.3 | 12.7 | 12.5 | 12.8 | Low test rate - Low CoV |
| Wound/Skin MCS | Miscellaneous | 5.0 | 27.1 | 26.9 | 27.2 | Low test rate - High CoV |
| Spirometry | Miscellaneous | 4.6 | 19.0 | 18.9 | 19.1 | Low test rate - High CoV |
| Hearing test | Miscellaneous | 4.2 | 51.6 | 51.4 | 51.7 | Low test rate - High CoV |
| Immunoglobulins | Blood | 4.0 | 22.2 | 22.1 | 22.3 | Low test rate - High CoV |
| CXR | Imaging | 3.8 | 17.9 | 17.8 | 18.1 | Low test rate - High CoV |
| Enteric virus screen | Miscellaneous | 3.8 | 26.2 | 26.1 | 26.4 | Low test rate - High CoV |
| Stool OCP | Miscellaneous | 2.4 | 20.4 | 20.3 | 20.5 | Low test rate - High CoV |
| Specific IgE | Blood | 2.4 | 16.5 | 16.4 | 16.7 | Low test rate - High CoV |
| Helicobacter pylori | Miscellaneous | 2.1 | 37.0 | 36.8 | 37.1 | Low test rate - High CoV |
| Faecal calprotectin | Miscellaneous | 1.8 | 10.4 | 10.3 | 10.6 | Low test rate - Low CoV |
| Monospot | Blood | 1.6 | 31.7 | 31.5 | 31.8 | Low test rate - High CoV |
| US abdomen | Imaging | 1.6 | 13.1 | 13.0 | 13.3 | Low test rate - Low CoV |
| US renal | Imaging | 0.8 | 14.1 | 14.0 | 14.3 | Low test rate - Low CoV |
| MRI head | Imaging | 0.7 | 22.8 | 22.6 | 22.9 | Low test rate - High CoV |
| CT head | Imaging | 0.2 | 16.5 | 16.4 | 16.6 | Low test rate - High CoV |
| FeNO | Miscellaneous | 0.2 | 123.7 | 123.6 | 123.9 | Low test rate - High CoV |

*High and Low in relation to test rate and variability are relative to the median test rate of 6.9 tests/1,000 child-years and median CoV of 16.5%

Abbreviations: CRP – C reactive protein; CT – computed tomography; CXR – Chest X-ray; ECG – Electrocardiogram; ESR – Erythrocyte sedimentation rate; FBC – Full blood count; FeNO – Fractional exhaled nitric oxide; LFT – Liver function test; MCS – Microscopy, culture, sensitivities; MRI – Magnetic resonance imaging; NOS – Not otherwise specified; OCP – Ova/cysts/parasites; TFT – Thyroid function test; US – Ultrasound
